# Strengthening School Water, Sanitation and Hygiene (WASH) Programme Implementation: Evidence from Expert Consensus in Uasin Gishu County, Kenya

**DOI:** 10.64898/2026.04.14.26350916

**Authors:** Gladys Chepkorir Seroney, Ng’wena A.G. Magak, Gugu Gladness Mchunu

## Abstract

**Introduction:** Access to safe water, sanitation, and hygiene (WASH) in schools is critical for child health, learning, and gender equity. In Kenya, the Kenya School Health Policy and the Basic Education Act outline standards for school WASH; however, implementation remains uneven due to inadequate infrastructure, weak inter-sectoral coordination, and limited financing. This study aimed to identify priority components for strengthening school WASH implementation and generate policy-relevant recommendations based on expert consensus in Uasin Gishu County, Kenya.

**Methods and Results:** A Delphi technique consisting of two iterative rounds was used to reach expert consensus. In Round 1, 20 purposively selected experts including head teachers, county education officials, public health officers, water and public works officers, and NGO representatives participated in key informant interviews. Emergent themes informed development of a structured Round 2 questionnaire administered through CommCare online app. Quantitative data were analyzed using descriptive statistics (means, standard deviations, percentage agreement), while qualitative responses underwent thematic coding using NVivo 12.

Experts reached strong consensus on essential components required for strengthening school WASH implementation. Key priorities included clear governance structures, designated budget lines, inclusive infrastructure, menstrual hygiene management (MHM), curriculum integration, sustained capacity building, and systematic monitoring. Multi-sectoral collaboration and recognition of best-performing schools were also emphasized as important motivators for compliance and sustainability. Equity considerations particularly the need for disability-friendly facilities and school-community outreach were highlighted as critical. Agreement levels ranged from 74% to 100%, with most items scoring mean values between 4.5 and 4.8 on a 5-point Likert scale, indicating strong consensus among experts.

**Conclusion:** strengthening implementation of school WASH in Kenya requires coordinated governance, predictable funding, reliable water systems, inclusive sanitation, strengthened MHM, and consistent monitoring beyond infrastructure investment alone. Integrating these expert-validated priorities within existing national policies offers a practical pathway to improving learner health, reducing absenteeism especially among girls and promoting equitable educational outcomes.

## Introduction

Access to safe water, adequate sanitation, and hygiene (WASH) services in schools is fundamental to promoting health, educational attainment, and gender equity among learners. In Kenya, the implementation of school WASH programs has gained increasing attention due to its direct impact on school attendance, learning outcomes, and the well-being of children, particularly girls and those with disabilities (1, 2)

Despite the existence of national policies such as the Kenya School Health Policy and the Basic Education Act, many schools continue to face challenges related to inadequate infrastructure, poor maintenance, and limited funding (3, 4). The lack of dedicated budget lines for WASH activities has resulted in fragmented implementation, with schools often relying on donor support or ad hoc community contributions (5).

Effective school WASH policies must address these gaps by establishing clear governance structures, ensuring sustainable financing, and promoting inclusive infrastructure. This includes the provision of clean water sources, modern sanitation facilities, and hand washing stations that are accessible to all learners, including those with physical disabilities (6). Moreover, menstrual hygiene management (MHM) must be integrated into WASH policies to support girls’ education and dignity. Studies have shown that inadequate MHM facilities contribute to absenteeism and dropout among adolescent girls (7, 8).

The inclusion of WASH education in the school curriculum is also critical. Embedding hygiene topics in science and life skills subjects, alongside extracurricular activities such as health clubs and commemorative events, fosters behavior change and community engagement (9, 10). Regular monitoring and supervision by public health officers and education inspectors are essential to ensure compliance and continuous improvement (11). In summary, a robust school WASH policy in Kenya must be multi-sectoral, adequately funded, inclusive, and aligned with national education and health strategies. It should empower schools to deliver safe, healthy, and equitable learning environments for all children. This study aimed to identify key elements for strengthening implementation of school WASH programs and propose policy recommendations based on expert consultations in Uasin Gishu County, Kenya.

## Materials and Methods

### Research Design

Consensus method using the Delphi techniques was used in the study. Delphi technique is a methodology for structuring a group communication process so that the process is effective in allowing a group of individuals (experts), to collectively address complex problems with a view to providing solutions (12).

The researcher engaged experts in school WASH program in various sub-counties and the county, school head teachers also participate as part of the expert panel responding to the two stages of the consensus Delphi process (Figure 1). Delphi technique was appropriate because each expert could freely provide their opinions privately, without meeting physically in round one of Delphi method and that Delphi technique is an appropriate method to reach consensus (13).

**Figure 1:**
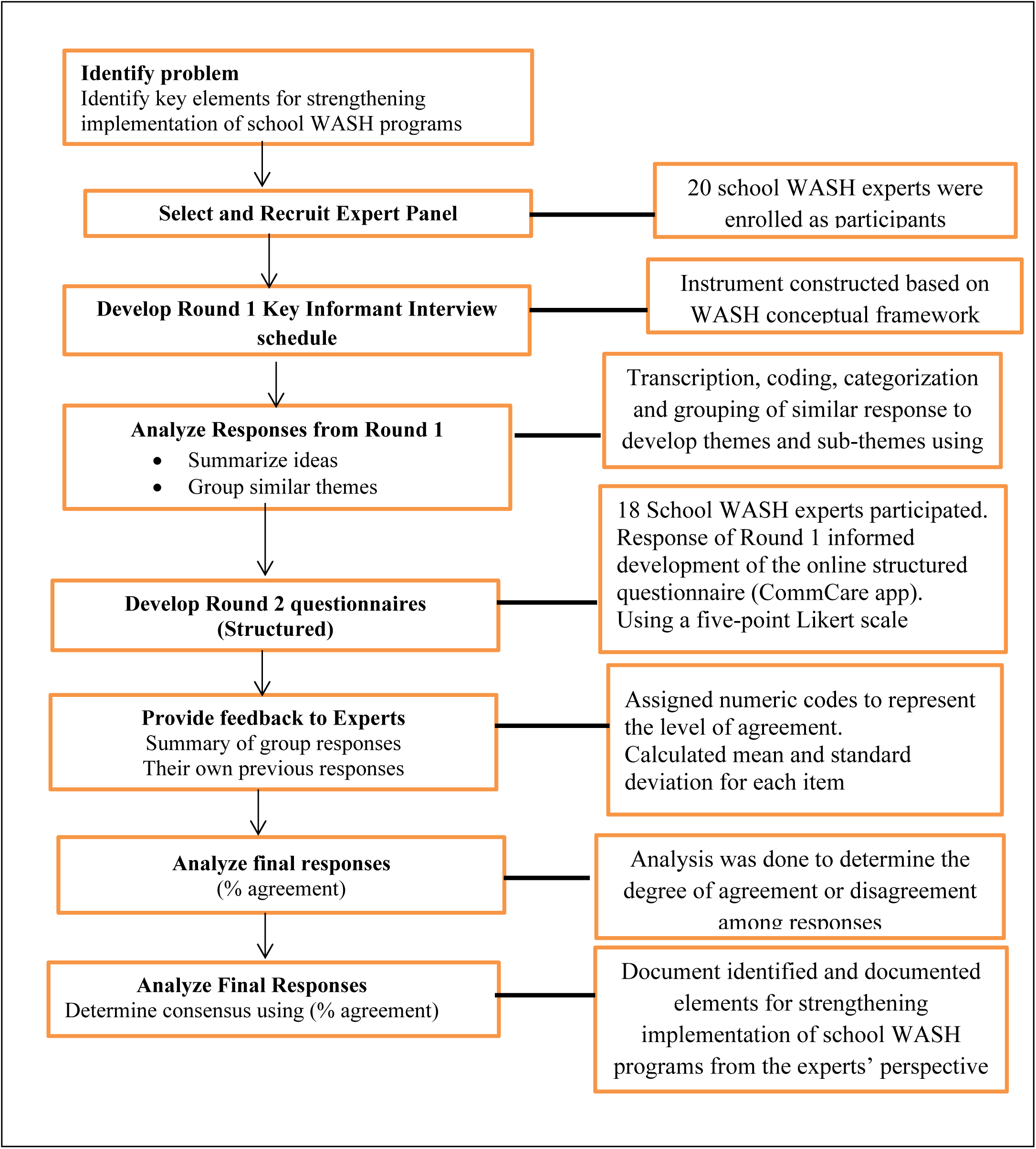
Flowchart of Delphi Technique process. Delphi Technique process ref: (14)

### Sampling

#### Selection of the panel of experts (key informants)

An expert is defined as a person with relevant knowledge of and experience in school WASH program. These experts were identified by their roles and responsibilities in school WASH as outlined in the Kenya School Health implementation guidelines (3). The following categories of experts were identified: the school head teachers, the County Educational directors, the schools’ Boards of Management (BOM), the county in-charge of water department, public health officers, the county public works officer, quality assurance and standards officers, and representatives of non-governmental organizations/ development partners participating in WASH activities in Uasin Gishu County.

### Sample size

County management officials’ in-charge of school health program in Uasin Gishu County, and any other organization participating in WASH activities in the county were purposively selected because the researcher was specifically interested in those who participate in WASH program. At the county and in each Sub-County there are specific and few personnel under the department of public health in-charge of WASH program.

It is recommended that 10-18 experts can be on a Delphi panel. However, Delphi group size does not depend on statistical power, but rather on group dynamics for arriving at consensus among experts (15). According to (16), sample size varies and depends on the research topic. For this study the researcher engaged only the personnel who are managing or engaged in WASH activities in the county, many of whom are employees of the government and are not many. Head teachers were also purposely sampled since they are the ones in-charge or managing the WASH program in schools

### Data collection methods and instruments

#### Delphi Round I

Identified experts were contacted in person and invited to participate. In Delphi round 1, the researcher conducted interviews using an interview schedule. The interview schedule was designed to gather information on policy and guidelines in place for executing the WASH program in primary school, planning and coordinating WASH program in schools, financing, budgeting monitoring and evaluation of school WASH program in the county and proposal on what needs to be done to improve WASH in schools.

The instrument was constructed based on the input component of the WASH conceptual framework for schools applied in this study and the appropriate research question. The Key Informants Interview (KII) was audio recorded; the interview took around 30-40 minutes. The interviewing of participants continued until no new information was gathered, that is when data collection stopped, this is referred to as the point of data saturation (17). Data collection for Delphi technique round 1 was collected from 1^st^ June 2025- 2^nd^ July 2025 following Ethical clearance from the Institutional Research Ethics Committee (IREC) of the Durban University of Technology in South Africa, ethical clearance was obtained on 28^th^ May,2025.

#### Delphi Round 2

Findings from Round 1 of Delphi technique informed the development of a structured questionnaire, which was used in a subsequent Delphi technique Round 2. The aim of the round was to give feedback to the experts, allow them to validate their responses to Round one and indicate their level of agreement or disagreement and re-rank their initial scores.

A self-administered online questionnaire was used to collect data from the experts using CommCare app that supported offline mobile data capture. Once data was collected and an internet connection is available, the data was automatically synced to the server. The experts were then asked to indicate their level of agreement or disagreement with the presented findings and re-rank their responses should they wish to do so. An Excel file was then generated and made available for download to facilitate data cleaning and further analysis.

All questions assessing agreement were measured using a five-point Likert scale, ranging from strongly disagrees to strongly agree. Each response was assigned a numeric code to represent the level of agreement: 1 for strongly disagree, 2 for disagree, 3 for neither agree nor disagree, 4 for agree, and 5 for strongly agree.

To evaluate overall agreement levels, the researcher calculated the mean and standard deviation for each item. In addition, the researcher considered responses of 4 (agree) and 5 (strongly agree) as indicating strong or “perfect” agreement. The proportion of respondents selecting either of these two options was computed to provide a summary measure of high agreement. Data collection for Delphi technique round to was conducted from 3^rd^ September to 10^th^ October,2025.

### Data analysis

#### Delphi Round 1

Audio-recorded interviews were transcribed verbatim into Microsoft Word. Transcripts were then imported into NVivo (version 12) for analysis. A coding framework was developed based on the study objectives and interview guide, and the researcher conducted deductive coding using NVivo to organize and categorize the data. Similar responses were grouped, and themes were developed by merging categories with related content.

#### Delphi Round 2

Data that was obtained from the second round of Delphi method was analyzed for the degree of agreement or disagreement in respondents’ (experts) responses. Data was thereafter summarized, identifying categories, and the obtained findings were used to finalize the development of the appropriate school WASH intervention that had emerged from the findings of the experts. Any major disagreements were resolved through consultation with the existing literature or concerned respondents to reach consensus

### Ethical approval and consent to participate

This study was conducted in accordance with the ethical principles outlined in the Declaration of Helsinki for research involving human participants. Ethical clearance was obtained from the Institutional Research Ethics Committee (IREC) of the Durban University of Technology, South Africa, and from the Ethical Review Committee of the University of Eastern Africa, Baraton, Kenya. Additional authorization to conduct the study in Kenya was granted by the Ministry of Education in Uasin Gishu County and the National Commission for Science, Technology and Innovation (NACOSTI). IREC Ethical Clearance number was (IREC 113/23)

Information sheet and letters of consent were shared with individuals requesting their participation in the Delphi study and the entire study. In these letters participants were informed that their participation was voluntary and they would withdraw at any stage of the study. All experts were over 18 years of age. Survey responses were anonymous as no personal identifiers or information was required from participating individuals. Participants were then asked to sign consent forms if they agreed to participate in the study.

## Results

### Characteristics of the school WASH experts

A total of 20 individuals signed up to participate in the Delphi process. The total number WHO participated in Key Informant Interviews (KIIs) formed Round 1 of the Delphi Technique. Table 1 presents the composition of the experts. They included: Sub-County Education directors, County officer in-charge of water department, County public works officer, public health officers, quality assurance and standards officers, constituency development funds officer, school head teachers and school board of Management. There was 90% response rate in Delphi Technique round 2 process, two (2) experts did not participate. The two experts were inaccessible since they had been transferred to other counties far from Uasin Gishu County.

**Table 1:** School WASH experts.

| **Key Informants categories** | **Allocated codes** | **Number** |
| --- | --- | --- |
| Sub-County Education directors | KII-EDU | 3 |
| County officer in-charge of water department | KII-ENG | 1 |
| County public works officer | KII-PWO | 1 |
| public health officers | KII-PHO | 3 |
| quality assurance and standards officers | KII-QSO | 3 |
| constituency development funds officer | KII-WPT | 1 |
| school head teachers | KII-SHT | 6 |
| school board of Management | KII-BOM | 2 |
|  | Total | 20 |

### Research results for Delphi technique round 1

The interviews focused on the implementation of school WASH program as well as the proposals on interventions to improve school WASH program in Uasin Gishu County, Kenya.

The results covered three overarching themes, namely (1) the personnel to be involved in implementing WASH program in schools, (2) what needs to be included in WASH policy guidelines in primary schools and (3) how WASH policy guidelines can be implemented in schools.

### Emerging themes

#### Theme 1: Experts view on personnel to be involved in WASH program in schools Multi-sectorial collaborations

As outlined in the National School Health Strategy Implementation Plan 2011-2015, Water Sanitation and Hygiene component of the school health program has many officers from several departments being engaged in school WASH, these include offers from various government ministries and other stakeholders as follows; ministry of education, ministry of public health and sanitation, Public works, ministry of health, water department, constituency development officer, Faith based organizations, non-governmental organizations, schools board of management, the school head teachers, Kenya Institute of Curriculum Development and many more.

> *Think everyone plays a role from the government agencies, to the… the people themselves, at the community level, to even our public health department. It’s a whole chain that needs to… they need to work together for the effectiveness (KII-ENG-01)*
>
> *Okay, for wash program, it’s a multi- sectorial thing, (shuffling noises) … it needs to be done by many, because if you work alone as a department, then we might not reach whatever is required. So, we need to have a forum of all the stakeholder, like I’d said, education should be there, health should be there, water and the environment, and the… all of us… everybody should be put on board, so that, we can achieve our… our target or whatever we want because if you work and do them in isolation then we might not achieve whatever we want (KII-PHO-02)*
>
> *It should be the responsibility of everybody who will have a supervisory role in our institutions, and it should not be limited to Ministry of Education. It should go beyond (KII-QSO-02)*.
>
> *Beyond the Ministry of Education, even within the Ministry of Education like I’ve seen I’m a quality assurance, we have the administrators and some county directors, theirs should not just be management alone. They should also be empowered to a level where when they visit institutions just approach the principal’s office or head teachers’ office, they should be able to move around and be able to raise those issues of WASH. There are those people who are working with the Teachers Service Commission, yeah it should not just be when they go to institutions, they are just looking at issues of teacher adequacy, discipline and all that, they should also be empowered so that they are also looking at issues in those institution where their teachers are working.(KII-QSO-02)*

### Partnership of both Public and private primary schools

It was observed that the stakeholders at school level were only from public schools and there was a suggestion to include private schools as stakeholders in school WASH programs since all school-going children either from public or private school have the right to access clean and safe water in adequate quantities and reasonable standards of sanitation.

> *Then I also think it’s good for us to mostly bring in the private entities as well, who could be having a lot more to offer, but they are not having properly laid down structures on how they can come in and help in that program KII-SHT-06*
>
> *is it disinterested, then there would be no sustainability, and that’s why most of the time, we talk about public private partnership, because now, we are the public, now private are the partners, and now only need to partner for sustainability. (KII-PHO-01)*

### Involvement of Community Health Extension Worker (CHEW) and the Reproductive health nurse in school WASH

Community Health Extension Worker (CHEW) and the Reproductive Health Nurse are members of health care system. Since (CHEW) works within the community, it was proposed that they be involved in WASH activities in schools within their jurisdiction and the reproductive health personnel to participate in MHM in schools

> *The CHEW is a public health and the nurse, so that when we do the… all the information when we go to schools, if I talk about WASH, a nurse will also talk about RH, because RH with MHM is related (KII-PHO-03)*

### Theme 2: Element for inclusion in School WASH policy and guidelines

#### Clear guide on coordination of school WASH activities

The experts felt that even with the governance structure outlines in the school health policy, coordination of WASH activities was not synchronized well from the top to the bottom. It was recommended that there be clear coordination of activities of school health and WASH program citing the example of how activities were conducted in schools during Covid 19 pandemic from the central government and county government.

> *You know the time of COVID, there was a very nice system and structure which was done, and you found that during that time the year 2020… 2020/21 – 2022, water, hygiene and sanitation in schools was perfect because it was well coordinated.(KII-SHT-02)*

#### Funding of WASH program by the government

It emerged that the funding of WASH components in schools was not clear. The school health policy, strategies and guidelines did not indicate where funds are coming from or if the government was funding all WASH activities.

> *I think what should be included is there would have been a policy that talks about funding by the government, rather than leaving it open, whereby the government can decide to fund or not fund. So, if there would have been, then it would have been so many steps ahead, because it would have been part of the policy… yes. Just like the way drugs are, you know the drugs must be bought to all this and this (KII-PHO-01)*

#### Outsourcing of school WASH services

For WASH services to be strengthened, it is proposed that school WASH policy and guidelines included information on outsourcing of WASH services to schools, as is reported outsourcing could improve WASH services and ease the burden from teachers so that they can concentrate in teaching. Some of the services recommended for out-sourcing include refuse and other waste collection and disposal, water and water resources provision and maintenance, Education to school teachers and pupils on WASH and especial Menstrual Hygiene management to girls who reach the age of receiving their menses:

> *May be if there was a policy that every institution, every school should have water from the source that is ELDOWAS for us in Eldoret, it would help for sanitation or drinking water (KII-SHT-05)*
>
> *We have companies that will come maybe once in a week or twice and make sure that the compound is clean. What will be left there is for the management now to make sure that that cleanliness is maintained. Until the next time when they’ll come back (KII-SHT-03)*
>
> *Because teachers cannot concentrate on that, and they are employed to teach. They should have people who are specialists in waters and sanitation (KII-SHT-02)*
>
> *I think sanitation in toilets, the schools should hire someone who is… who is paid. Yes, it should not be the students alone. The students can be taught on making the toilet clean, but there should be somebody who is in charge to make sure that the toilets are always clean (KII-BOM-01)*.
>
> *Other schools have contracted companies to provide training to teachers on water treatment and other schools have companies contracted to provide bins for disposing sanitary towels and collecting filled bins*.

#### Government to provide Water to all private and public schools

Water being an essential commodity to all school-going children it was suggested that, the provision of water and maintenance of its resources should be done to both public and private schools and this should be the prerogative of the government.

> *think it has to start from the top by government and in fact even government itself relooking into the whole program of water and hygiene programs in our schools. In that to sort out these problems we have for example, where schools are not so well connected to the water and the services, instead of saying a school for example should separately pay for its water services to get them I thought it should be a declaration of government, that any learning institution whether it’s a private one or a public one should have access*
>
> *to water and then they can devise ways of getting the money they needed to actually sustain the programs instead of saying if the school for example, is not able at a particular point you know to pay up for those services then they are disconnected. I think that can be a better approach (KII-SHT-06)*

### All schools to have sunken boreholes

To ensure constant supply of water for hand washing, drinking and other uses it was recommended that all schools have sunken boreholes and connected to solar system so that schools could reduce the cost of pumping water from the borehole and reduce the cost of electricity bills.

> *We have a well but that one is not sufficient. Sometimes when it is dry season, we have to get enough water, yeah. So, the water should actually be drilled, yeah in school in all the schools KII-SHT-04*
>
> *If it will be easier, I hope that every school should have borehole and solar power, where each school will be having its own water in the school* KII-BOM-02

### All schools to have modern toilets with septic tanks or connected to sewerage systems

During interviews, it was reported that water table in Uasin Gishu county is a little high and so during the rainy season water levels in the pit latrines rises and the latrines cannot be used due to lack of safety brought by increased tendencies of toilets collapsing, danger of splashing during use and fear of outbreak of diseases due to overflow of human waste and contamination of water and environment. It is from this background that there were suggestions that all schools should have modern flush toilets connected to septic tanks or to the sewerage system depending on their location.

> *Okay, on sanitation I would say that the government also should provide modern toilets, the flash toilets rather than the pit toilets. Especially in this area, because there is a lot of water. In fact, I recently requested a head teacher that we make a report so that we recommend the flash toilets. For this school, because this is a place where the water level is… (crosstalk) So if we can have, have flash toilets, then we have a septic tank or a bio digester. That could be better. Yeah. And it could be safe for the children, but because of funds now we don’t know what to do (KII-SHT-04)*
>
> *If every school get its own water, source of water, my suggestion is that we should not be using pit latrines, nowadays, we should be using modern toilets whereby we should use digesters so that we avoid each and every year where toilets collapse, maybe some pupils are injured. If we use modern technology, modern toilets, the only challenge is that it requires water (KII-BOM- 02)*

### Increase frequency of school inspection visits

It was reported that the county government has sub-county quality and standards officers inspecting schools to maintain standards through the Ministry of Education. When it comes to inspection and maintenance of WASH infrastructure, the respondents recommend that it is necessary to include in the WASH guidelines the number of visits an officer needs to make to a school within a given time and also increase the frequency of school inspection visits.

*Then thirdly, they should have an inspection done every quarter. They should send the… government should send inspectors in schools to ensure that the hygiene, water and sanitation is in place (KII-BOM-01)*

> *They come and inspect and also advise the school institution on how to improve the areas they have not yet improved and… and probably visit severally to make sure that whatever is being discussed is implemented because the moment they come we discuss and then they disappear, we shall also forget about it. But the moment they come regularly, we shall not be knowing that in a week time or two weeks’ time, these people will come (KII-SHT- 03)*

### Adding WASH content to school curriculum

Water, sanitation and hygiene in primary schools in Kenya is included in the curriculum, the three components are covered as separate entities and taught in different classes. It is suggested that WASH education content be increased in primary school curriculum and maybe for it to be taught as an independent subject.

> *Yes, it’s true. I think sensitization should be included even in the education curriculum like every class every grade should have a lesson just about water and sanitation and hygiene and if possible, every month they should be just a class about that. (KII-SHT-01)*
>
> *So, we are proposing to the government so that maybe, they would put it in guidelines that when they are coming up with the curriculum, they should ensure that WASH comes out of the subject (KII-SHT-04)*

### Increase budgetary allocation to school WASH

All schools in the county have a budget allocation from the county government for running schools activities and each learner has a vote head, the head teachers indicated that the vote head was very minimal. The money was not adequate to provide and maintain WASH facilities and other activities. Since WASH activities were very essential to the learners and required a lot of resources, there was a proposal that WASH activities be allocated a vote and not to be included in a general vote for the school.

> *I’m not saying that the votes is there, that if the government can really provide and specify that this water… this… this money is for water and sanitation (KII-SHT-01)*

*No, it’s not like for example what they are giving us on EWC which is electricity, water and conservancy, very little, as little as 108 shillings per term per child. So, it doesn’t even pay one month’s bill, yes. So, say repairs, say payment of workers and day to day consumables, it’s hectic. It is God taking care of…(KII-SHT- 02)*

> *It’s not possible. It may not be possible until and when, the National government provides enough capitation… yes (KII-SHT-02)*

#### Ensure availability and adequate supply of sanitary towels for menstrual hygiene in schools

It emergent that there was need for coordination in the distribution of the sanitary towels to schools for Menstrual Health Management and planning to mobilize resources to support menstrual hygiene activities in schools. The respondents indicated that the supply of the menstrual products was inconsistent and inadequate.

> *, and we do supply them with sanitary towels. That is from the ministry, though this supplies normally comes maybe, once a year, but at times they are not enough for these girls. So, we end up maybe, sourcing from other sources KII-SHT-01*
>
> *What the ministry can do or the county government, is to make sure that we have constant supply of water, constant supply of the sanitary pads, and maybe, once…KII-SHT-01*

#### Availing menstrual waste disposal bins and construction of changing rooms for girls

Some schools had waste disposal bins in their schools while others especially public primary schools did not have menstrual waste disposal bins and separate changing spaces for girls. It is recommended that the government should provide sanitary bins and enough sanitary towels to school girls.

> *we have… we have after installing the bins, they have now understood, that when it is full, they are able to report. The idea of reporting is there (KII-STH-02)*.
>
> *Everything is okay on that side. Where to put sanitary towers and those kinds of things (KII-SHT-03)*.
>
> *Yes… yes. The government should also provide those enough sanitary pads for the girls. The government should provide… what do we call… like this one for Rentokil, yeah, where they can dispose, yeah, the pads after use. So, all the girl’s toilet should have (KII-SHT-04)*
>
> *Actually, we don’t have. But remember that, this school was started long time ago, it is not a modern one, but we believe that if we are to be built another toilet, another washroom, it will have it (KII-SHT-01)*

#### Conducting outreach and sensitization activities on MHM in schools

The experts also proposed the need for outreach and sensitization activities in schools on Menstrual Health Management

#### Adaptability of all schools with facilities to meet the needs of children with special needs and disability

A component of special needs, disability and rehabilitation is included in the school health policy and the adaptation of school environment to accommodate children with mobility related disabilities. It emerged from the interviews that there were some categories of schools that were for children with mobility related disabilities, these schools is where there were recommendations on installation of WASH facilities.

> *Okay we have categories of schools. We have the special schools. So, in, in case of the special schools, we always, it’s a mandatory that the facilities are friendly to the learners with disabilities. Yes. So, they’re, they’re able to access (KII-EDU-01)*

It was indicated that many schools lacked disability friendly environment thus hindering admission and retention of such learners to school. Schools lack ramps, special toilets and special handles for opening hand washing points and hand washing positions may not be out of reach to the learners with physical challenges. It is suggested that all schools and new ones should have provisions for children with mobility since the government is advocating for integration in schools for all learners and social participation.

> *Yes, all the classes in the areas of water points should be friendly to those children with challenges, yes. The ramp should be made around the water points, classes, toilets, yes (KII-SHT-04)*
>
> *For the case of integrated schools. We, as much as possible, ensure that such facilities are provided. They may not be adequate for now. But that is where, where we are heading to. Yes (KII-EDU-01)*

#### Addition of technical support

The informants felt that there was need for additional technical support in implementing, operation and maintenance of school WASH infrastructure.

> *whatever guidelines are there suffices. If they could be any other thing to be added, that could be the technical aspect of it, the technical people should come in now but for us that suffices, I think it’s adequate… it’s adequate… it’s adequate (KII-QSO-02)*

#### Theme 3: Ways of implementing school WASH Policy and guidelines Commemoration of WASH days in schools

It emerged that several strategies have been employed to improve access to school WASH, create awareness and promote good hygiene habits among school going children. Some of these activities that can conducted in schools include commemorate WASH days which include Global hand washing day, menstrual health management day, world toilet day and world environmental day. One informant had this to share

> *We also have World Hygiene Day. On that World Hygiene Day, they can do it in our schools, whereby they create awareness, they provide sanitary pads, they provide the soaps, they provide anything that we feel is related to the sanitation*. KII SHT 01
>
> *For sensitization. So, they make it sort of a policy that on those days that are essentially related to water, sanitation is commemorated in schools KII-SHT-01*

#### Parents and community engagement

It was mentioned that for WASH activities to succeed, communities where children came from must be engage in WASH activities though funding school WASH activities and they should also be sensitized on hygiene issues, this was reported.

> *Since most of them require funding, I think it is upon all the stakeholders, particularly government in the sight of parents and BOM we should also chip in, community will also have its own role to chip in so that students or pupils should also participate in implementing the guidelines KII BOM 02*
>
> *It is good that in CBC hygiene is, there is a subject of hygiene. Hygiene should also be taught in the community, whereby if students or pupils are taught in school, they will also… we encourage them to teach their parents or the community so that we have hygiene at home and we have hygiene at school, not only in our schools KII BOM 02*

#### Capacity building of teachers and health promotion on WASH in mainstream media

Though WASH is taught in the school curriculum some informants felt there was a need to educate the teacher and learners on WASH by the county government, others felt that sensitization could be done through mass media. Informants indicated that;

> *The county government, together with the national government, at times it is good that they send their people to come and educate us about sanitation and water and all those*
>
> *things. Occasionally, maybe it can be gradually or irregularly so, that we don’t forget about it (KII SHT- 05)*
>
> *People should be sensitized through the radios, through the… through the… (someone talking in the background) through the media for everybody to get to know that it is very important for us to make sure that where we live in, is safe and the sanitation is good (KII- SHT- 01)*.

#### Regular and routine supervisory of WASH in schools

It emerged that officers conducting supervision maned large areas and were expected to conduct supervision of the schools within a given time frame.

> *…if I give an example of the quality assurance officers here in this sub county Ministry of Education, and then you’d be looking at the institutions of the numbers, like secondary schools, then you look at the primary schools, you’re talking about 200 plus institutions. So, with that kind of a number definitely you’re the one who is doing that and maybe it’s a requirement that you visit a school within a period of three years, so you might leave this place and go to another sub county minus visiting quite a number of those schools (KII-QSO-02)*
>
> *They should be employing people to be monitoring on day to day or routine management. You should have routine management, so that they look when they to make sure that water, hygiene and sanitation is updated KII-STH-0 SOSIANI*

The head teachers recommend that school supervision of WASH facilities ought to be conducted regularly, feedback following supervision would be timely given and reports from the supervision ought to be handed over for action.

> *Now that one is under public health, they could be coming all the time and making sure that the sanitation is ok, write reports, take the reports to the ministry of education, may be somethings would be taken in seriously. But you see they come only ones may be when we call them, example we when we want a toilet or we want to construct a building, you find that that is when they come, we need a lot of regular, ok they come but they should come specifically for sanitation and also get a report and present that report to other offices so that action should be taken. Check on the water, the learners, kitchen and everything and give a report. I think something can be done(KII- SHT- 05)*
>
> *We expect the public health officer to make sure that the water points are to check whether it is it has any hazard or health hazard and to give advice, yes. And to do the testing of water KII- SHT- 04*.
>
> *…who are dealing with the issue of water, they should be regular visitors to those institutions. We have the public health they should not come in just to do the report in our schools (people talking in the background) but the become a routine to them. They should have a routine to go to through institutions on a regular basis to have a look at that (KII-QSO-02)*.

#### Additional of supervisory personnel

An informants reported that some officers did not do due diligence in supervision of school WASH and could just sign the visitors’ book. The officers were urged to be diligent in their work. It was reported that the officers deployed for the work were also few, one school head teacher said

> *Quality and assurance standards for officers in the county are very few. They are not enough. And then. They should be doing their work diligently because we have seen in some schools they just go there; they sign the Visitor’s book in the office and they leave. They don’t visit those areas, especially the toilets and any other area that needs improvement for example. So, they should do their work diligently and we should have enough of those officers KII SHT 01*.

#### Inspection and monitoring of funded facilities in schools

It was noted that sometimes funds could be given towards WASH activities in schools and follow up on how the money was utilized and evidence of activities done was not followed up by the relevant authorities. It is proposed that the follow-up of funds be made for accountability purposes.

> *They should also make a follow up to make sure that the same money has been used in the right way KII- SHT- 01*

#### Recognizing and awarding best performing schools on WASH

It emerged that recognition and awarding schools with the best WASH facilities or infrastructure could motivate school management and learners to engage in WASH activities hence improving their health states. One informant had this to share

> *Yeah, they can come in big time. Like I’ve said, first, they should be very, very present. And then number two, they can come in to… to do appraisals, for example, where certain institutions have done so well in the implementation of those policies, they can come in, recognize those institutions, award where necessary and also identify which ones are struggling and support them apart from identifying those who are… who are the stumbling blocks as well in a implementation of the same policies KII SHT 06*

#### Appointment of schoolteacher/s or a department to coordinate school WASH

In some schools, teachers have been appointed to be responsible for WASH activities, in other schools the teacher on duty takes responsibility while other schools had there is a department constituted to manage WASH activities.

> *Teacher… Teachers On Duty, Heads of Departments… yes. Those are major people who are majorly concerned being coordinated by one of my deputies (KII-SHT-02)*

#### Board of Management to support maintenance of WASH facilities

It emerged that some of the challenges that hindered access to WASH facilities was vandalism of WASH facilities, poor maintenance and misuse of water resources. It was therefore proposed that the BOM support the maintenance of WASH facilities in their schools.

> *The boards of management. Yeah. The boards of management. If, if they can up, their role in ensuring the same, I think we’ll be able to address that challenge (KII-QSO-03)*.

#### Research results for Delphi technique round 2

There was 90% response rate since 2 experts that participated in Delphi Technique round 1 did not participate. From the results, the experts generally showed high levels of agreement. The mean scores ranged for all items ranged between 4.5 and 4.8 indicating a tendency toward agreement across the board. Standard deviations were relatively low, suggesting limited variability in responses.

When examining the proportion of respondents who selected either agree or strongly agree, a substantial majority demonstrated perfect agreement on key items. Similar agreement patterns were observed across all items, highlighting consistent support for the constructs measured.

Table 2 provided data on level of agreements of structured questionnaire responses of experts on ways to make school WASH strengthened.

**Table 2:** Delphi Technique Round 2.

| Item | Meansd | Agree |
| --- | --- | --- |
| **Personnel to be involved in the implementation of WASH program in primary schools** |  |  |
| Community health extension worker chew needs to be involved in school WASH | 4.6 (SD 0.5) | 100% |
| For WASH in schools to be a success there must be collaboration among different stakeholders including the school board, County and health coordinator | 4.9 (SD 0.3) | 100% |
| Reproductive health nurse needs to be involved in school WASH | 4.4 (SD 0.8) | 95% |
| The government to involve public and private primary schools in wash program | 5 (SD 0) | 95% |
| **Interventions to be included in the WASH policy guidelines in primary schools** |  |  |
| Item | Meansd | Agree |
| Addition of wash content to school curriculum | 4.6 (SD 0.5) | 100% |
| Deployment of more quality assurance and standards officers and public heal | 4.7 (SD 0.6) | 95% |
| Inclusion of multiple health stakeholders on school wash program | 4.8 (SD 0.4) | 100% |
| Increase budgetary allocation to school wash | 4.7 (SD 0.5) | 100% |
| Increase frequency of school inspection visits | 4.8 (SD 0.4) | 100% |
| Need for partner support in implementation of the wash program in schools | 4.6 (SD 0.5) | 100% |
| Need for separate budget and funding for wash in schools | 4.5 (SD 1) | 95% |
| Newly established schools should be integrated schools -have friendly facilities | 4.9 (SD 0.3) | 100% |
| Outsourcing the maintenance of wash facilities in schools | 3.9 (SD 1) | 74% |
| The governments to fund school wash programs | 4.7 (SD 0.5) | 100% |
| *Water interventions* |  |  |
| Item | Meansd | Agree |
| All schools to have sunken boreholes and water storage tanks | 4.4 (SD 0.8) | 89% |
| Government to provide water to both public and private schools within them | 4.5 (SD 0.5) | 100% |
| Water analysis - routine testing of the quality of drinking water sources; physical, chemical, and microbiological contaminants | 4.7 (SD 0.5) | 100% |
| *Interventions on hygiene promotion among learners and teachers in the school* |  |  |
| *Item* | *Meansd* | *Agree* |
| Access to education material on wash for students both in schools and outside | 4.7 (SD 0.6) | 95% |
| Observe commemoration of important WASH days; world hand washing day, world toilet day | 4.7 (SD 0.5) | 100% |
| Introduction of health clubs and talking messages on wash in schools | 4.8 (SD 0.4) | 100% |
| Learners taken through practical on how to promote wash activities. | 4.7 (SD 0.5) | 100% |
| Outreach and sensitization activities in schools. | 4.7 (SD 0.5) | 100% |
| There is need for peer education on wash in schools. | 4.8 (SD 0.4) | 100% |
| *Menstrual health management* |  |  |
| Item | Meansd | Agree |
| All schools should have menstrual waste disposal bins | 4.8 (SD 0.4) | 100% |
| Changing rooms for girls during menstruation be constructed in every school | 4.8 (SD 0.4) | 100% |
| Government to ensure availability and adequate supply of sanitary towels | 4.8 (SD 0.4) | 100% |
| Plan and carry out world menstrual day | 4.7 (SD 0.5) | 100% |
| Services for collection and disposal of sanitary towels be outsourced | 4.5 (SD 0.5) | 100% |
| *Access to school WASH facilities by learners with disabilities* |  |  |
| Item | Meansd | Agree |
| All newly established schools to have ramp to easy movement of learners with disabilities | 4.8 (SD 0.4) | 100% |
| All newly established schools to have sanitation facilities appropriate for learners with disabilities | 4.9 (SD 0.2) | 100% |
| Taps height consideration and models of opening and closing the taps to be accommodative to learners with disabilities | 4.7 (SD 0.5) | 100% |
| *Sanitation facilities in schools* |  |  |
| Item | Meansd | Agree |
| All schools should have flush toilets connected to septic tank for emptying when needed or the sewerage system | 4.7 (SD 0.5) | 100% |
| Schools to have staff employed specifically to ensure cleanliness of the sanitation facilities and school environment | 4.6 (SD 0.6) | 95% |
| **Ways of implementing WASH policy guidelines in schools** |  |  |
| Item | Meansd | Agree |
| Best performing schools on WASH should be recognized and awarded | 4.6 (SD 0.5) | 100% |
| Government to engage both private and public schools' stakeholders on wash | 4.7 (SD 0.5) | 100% |
| Head teacher to appoint teachers or departments to coordinate school WASH | 4.8 (SD 0.4) | 100% |
| Head teachers and teachers need capacity building on WASH program in school | 4.7 (SD 0.5) | 100% |
| There is need for follow-up of funded facilities in schools to ensure completion. | 4.6 (SD 0.5) | 100% |
| Best performing schools on WASH should be recognized and awarded | 4.6 (SD 0.5) | 100% |
| Government to engage both private and public schools' stakeholders on wash | 4.7 (SD 0.5) | 100% |
| Head teacher to appoint teachers or departments to coordinate school WASH | 4.8 (SD 0.4) | 100% |
| Head teachers and teachers need capacity building on WASH program in school | 4.7 (SD 0.5) | 100% |
| There is need for follow-up of funded facilities in schools to ensure completion. | 4.6 (SD 0.5) | 100% |

#### Round 2 Delphi technique results, Experts consensus on strengthening school WASH program implementation in Uasin Gishu County Kenya

## Discussion

### Overview of principal findings

This Delphi study produced a strong, practice-oriented consensus on what is required to strengthen the implementation school WASH program in Uasin Gishu County. Experts agreed on the need for multi-sectoral governance; dedicated, ring-fenced financing; reliable water supply; upgraded, inclusive sanitation; robust menstrual health management (MHM) systems; and strengthened supervision and accountability. These priorities echo Kenya’s policy intentions but expose persistent implementation gaps that limit equity and sustainability of school WASH services, particularly for girls and learners with disabilities (18–20).

### Interpreting the expert consensus in light of policy and evidence

Quantitatively, Round-2 ratings were clustered at high agreement (means 4.5–4.8 on a 5-point scale, low SDs), with 95–100% endorsement on core items such as multi-sectoral collaboration, disability-inclusive designs, curriculum integration of hygiene, increased inspection frequency, and dedicated budget lines. This pattern was consistent with international evidence that infrastructure alone is insufficient without strong governance, monitoring, and recurrent financing that cover operation and maintenance, consumables (soap, water treatment, cleaning supplies), and MHM commodities (5, 20).

### Governance, roles, and stakeholder engagement

Experts called for clear vertical coordination from national to school level, citing the highly organized inter-governmental approach observed during COVID-19 as proof of what coordination can achieve in the short run. They endorsed a broadened constellation of actors education, health, water, public works, quality assurance, boards of management, CHEWs, reproductive health nurses, and development partners with explicit roles and shared accountability. Inclusion of private schools was emphasized to avoid dual standards in access and to align with universal service principles embedded in Kenya’s Basic Education Act and School Health Policy (4, 18, 20).

### Financing and sustainability

A recurring thread across interviews and consensus ratings was the inadequacy and fragmentation of current financing. Experts recommended creating a dedicated WASH budget line at school and county levels, rather than including WASH under broad votes that are quickly depleted by utilities and emergencies. This reflects global analyses that underfunding and lack of recurrent O&M budgets undermine functionality and service levels, even where infrastructure exists (5, 21).

Outsourcing selected services e.g., desludging, solid waste and menstrual waste management, periodic deep-cleaning, and targeted training was strongly supported to improve reliability and relieve teachers of technical burdens, consistent with comparative evidence showing efficiency gains when specialized providers are engaged (8, 22).

### Reliable water supply

The panel strongly favored school-based boreholes with solarized pumping and adequate storage to overcome intermittency and billing shocks from utility networks. This reflects experience in many East and West African school systems where boreholes, when safely sited and maintained, stabilize hand washing, drinking, and cleaning, and reduce absenteeism especially for girls during menstruation. Experts also underscored routine water quality testing to ensure safety for consumption and hygiene (23, 24).

### Sanitation infrastructure and service models

Given high water tables in parts of Uasin Gishu and recurrent structural failures of pit latrines, experts prioritized transition to improved, water-flush solutions connected to septic tanks or sewer lines, or to water-saving, waterborne alternatives where feasible. This is consistent with rights-based arguments that unsafe pits compromise dignity and safety, and with emerging models that drastically reduce per-capita flushing volumes while improving cleanliness and durability (25, 26).The consensus further endorsed employing paid staff responsible for routine cleanliness to ensure daily service levels are maintained.

### Menstrual health management (MHM)

The study highlights MHM as both a health and equity imperative. Experts reported inconsistent pad supplies, limited disposal options, and inadequate privacy for changing, and they endorsed comprehensive measures: reliable provision of menstrual products, disposal bins in girls’ toilets, gender-sensitive changing rooms, and contracted services for safe waste handling. This aligns with recent global monitoring that documents large gaps in MHM education, facilities, and product availability in low-resource settings, and with evidence linking improved MHM to attendance and psychosocial outcomes (1, 7, 8).

### Disability inclusion and universal design

Experts strongly supported making all new schools and progressively existing schools disability-friendly, citing ramps, accessible toilet compartments, handrails, and tap heights and mechanisms suited to a range of mobility and grip needs. This echoes SDG commitments and Kenya’s policy framing on inclusion, and responds to well-documented barriers that prevent learners with disabilities from safely accessing WASH spaces and from remaining in school (6, 27–29)

### Health education, curriculum integration, and behavior change

Although hygiene topics are included in the curriculum, experts emphasized the need to deepen curricular content, reinforce learning through school health clubs, and leverage commemorative WASH days for experiential learning and community engagement. They also supported sustained teacher capacity-building and use of mass media for wider health promotion. These recommendations are aligned with evidence that school-based health promotion must couple individual behavior approaches with supportive environments to translate knowledge into sustained practice (9, 10, 30).

### Monitoring, supervision, and accountability

The panel identified supervision as a cornerstone of implementation but noted staffing shortages, heavy workloads, and infrequent visits that limit problem-solving and feedback loops. Recommended actions included increasing the number of quality assurance and public health officers, establishing minimum inspection frequencies, standardizing tools and reporting pathways, and ensuring follow-up on funded works for accountability. Recognition and awards for best-performing schools were proposed to create positive competition and accelerate adoption of good practice an approach consistent with behavior change communication and performance-based incentives used in school health programs (31, 32).

### Community and parental engagement

Experts stressed that community buy-in and parental contributions financial, in-kind, and behavioral are indispensable. Extending WASH messages to households through learners and CHEWs can harmonize norms at home and school, creating a reinforcing environment for hygiene practices. This complements Kenya’s competency-based curriculum orientation and situates schools as hubs for community health promotion (18, 33).

### Strengths and limitations of the evidence

The Delphi approach enabled convergence across a diverse set of practitioners who manage or influence school WASH, producing practice-grounded recommendations. However, findings are context-specific to Uasin Gishu and reflect expert, rather than learner or parent, perspectives; future mixed-methods work could integrate student, caregiver, and facility-level performance data to triangulate and quantify impact pathways. Nonetheless, the panel’s high consensus and the close alignment with national and international guidance enhance transferability to similar Kenyan counties.

## Conclusion

Strengthening school WASH in Kenya cannot be achieved through infrastructure alone. The expert consensus from Uasin Gishu County points to a comprehensive implementation agenda: multi-sectoral governance with clearly defined roles; dedicated, ring-fenced financing for infrastructure, operation and maintenance, and MHM; reliable and safe water through school-level supply and testing; modern, disability-inclusive sanitation; reinforced health education and school-community engagement; and robust supervision with transparent follow-up and incentives for excellence. Operationalizing these priorities within existing policy frameworks provides a feasible pathway to improve learner health, reduce absenteeism particularly for girls and advance educational equity. Making these shifts routine rather than episodic requires political will, predictable finance, and disciplined accountability mechanisms that link standards to practice in every school.

## Recommendations

Based on the expert consensus and the interpretation of findings, several actionable recommendations are proposed to strengthen the implementation and sustainability of school WASH programs in Kenya.

The first recommendation is on strengthen multi-sectoral governance and coordination. The Ministry of Education, Ministry of Health, county governments, water departments, public works, CHEWs, reproductive health nurses, private schools, and development partners should adopt a more structured and harmonized coordination mechanism. Clear role delineation and reporting pathways are essential to ensure accountability and seamless implementation across levels.

Secondly, the government should establish dedicated budget lines for WASH to schools to address chronic underfunding. School WASH should receive a ring-fenced budget line at national, county, and school levels. This funding should specifically cover infrastructure development, routine operation and maintenance, hygiene supplies, and menstrual health management commodities to ensure continuity and program sustainability.

Thirdly, schools should prioritize sustainable water solutions, including drilling boreholes equipped with solar pumping systems and routine water quality testing. These measures will ensure consistent access to safe drinking and hand washing water, reducing disease risk and supporting daily school operations.

The ministry of Education should strengthen WASH Education, health promotion, and community engagement these can be achieved WASH content being expanded in the curriculum, supporting active school health clubs, commemorating school WASH days, and outreach activities. Engaging parents and communities, these will reinforce hygiene behaviors both at school and at home.

County governments should reinforce monitoring, inspection, and accountability mechanisms of School WASH by increasing the number of Quality Assurance and Standards Officers and Public Health Officers dedicated to WASH. Establishing minimum inspection frequencies and standard monitoring tools will improve compliance and timely feedback. Follow-up on WASH-related expenditures should be mandatory for transparency and accountability.

Lastly, it is recommended that Ministry of Education should promote recognition of WASH high-performing schools. Instituting awards for schools demonstrating excellence in WASH implementation can motivate improved performance, foster innovation, and promote peer learning across institutions. Such recognition can serve as an incentive to maintain high WASH standards.

## Data Availability

All data generated or analyzed during this study are included in this published article.

## Declarations

### Ethical approval and consent to participate

Information sheet and letters of consent were shared with individuals invited for participation in the Delphi study and the entire study. In these letters participants were informed that their participation was voluntary and they would withdraw at any stage of the study. All experts were over 18 years of age. Survey responses were anonymous as no personal identifiers or information was required from participating individuals. Participants were then asked to sign consent forms if they agreed to participate in the study.

### Consent for publication

The research paper does not contain any individual, personal data in any form. No consent for publication needed. Consent for publication was therefore not applicable.

### Availability of data and materials

All data generated or analyzed during this study are included in this published article.

### Authors’ contributions

GCS conceptualized the study and prepared the draft proposal under the supervision of GGM and MAGN.

GCS and GGM contributed to developing the background and designing the study. GCS prepared the manuscript, while GGM and MAGN critically reviewed it.

GCS accessed library resources and conducted data analysis.

All authors (GCS, GGM, MAGN) reviewed and approved the final manuscript.

## Acknowledgements

The authors would like to extend their appreciation to Durban University of Technology library for availing resources to be used for literature review and data analysis.

## Author’s information

Gladys Chepkorir Seroney is currently a PhD candidate in the faculty of health sciences in Durban University of Technology in South Africa. She is a lecturer in the Department of Community Health Nursing, School of Nursing, Maseno University, Kisumu, Kenya.

## Reference

1. United Nations Children’s Fund, World Health Organization. Progress on drinking water, sanitation and hygiene in schools 2015-2023: special focus on menstrual health 2024 [Available from: https://washdata.org/reports/jmp-2024-wash-schools.

2. Ministry of Education, Ministry of Health. Kenya School Health Implementation Guidelines. 2018 [Available from: https://repository.familyhealth.go.ke/xmlui/bitstream/handle/123456789/82/Implementation%20Guidelines%202nd%20Edition%202018.pdf?sequence=1&isAllowed=y.

3. Ministry of Education, Ministry of Health. Kenya School Health Policy. 20218. 2^nd^ Edition. https://nipfn.knbs.or.ke/download/kenya-school-health-policy-second-edition-2018/

4. Ministry of Education. The Basic Education Act 2013. Government of Kenya 2013 http://www.parliament.go.ke/sites/default/files/2017-05/BasicEducationActNo_14of2013.pdf

5. Deroo L, Walter E, Graham J. Monitoring and evaluation of WASH in schools programs: lessons from implementing organizations. Journal of Water, Sanitation and Hygiene for Development. 2015; 5(3):512–20. 10.2166/washdev.2015.026

6. Azupogo UW, Dassah E, Bisung E. Navigating water and sanitation environments in schools: Exploring health risk perceptions of children with physical disabilities using drawing. Wellbeing, Space and Society. 2025;8:100255. 10.1016/j.wss.2025.100255

7. Hennegan J, Montgomery P. Do menstrual hygiene management interventions improve education and psychosocial outcomes for women and girls in low and middle income countries? A systematic review. PloS one. 2016;11(2):e0146985. 10.1371/journal.pone.0146985

8. World Bank Group. The Enabling Environment for Menstrual Health and Hygiene: Case Study - Kenya. 2022 [Available from: https://openknowledge.worldbank.org/handle/10986/39725.

9. United Nations Children’s Fund. Water, Sanitation and Hygiene (WASH) in Schools Child friendly schools. A companion to the Child Friendly Schools Manual: UNICEF Division of Communication; 2012 [Available from: https://inee.org/sites/default/files/resources/CFS_WASH_E_web.pdf.

10. Morrish D, Neesam M. Trends in coverage of hygiene and disease prevention topics across national curriculum frameworks for primary science, physical education, and health. Prospects. 2021;51(1):363–81. 10.1007/s11125-020-09525-7

11. World Health Organization. Improving access to water, sanitation and hygiene can save 1.4 million lives per year, says new WHO report: World Health Organization 2023 [Available from: https://www.who.int/news/item/28-06-2023-improving-access-to-water--sanitation-and-hygiene-can-save-1.4-million-lives-per-year--says-new-who-report.

12. Fawole OI, Wyk JV, Adejimi AA, Akinsola OJ, Balogun O. Establishing consensus among interprofessional faculty on a genderbased violence curriculum in medical schools in Nigeria : a Delphi study. African Journal of Health Professions Education. 2018;10(2):106–13. https://journals.co.za/doi/abs/10.7196/AJHPE.2018.v10i2.988

13. Grutters JPC, Bouttell J, Abrishami P, Ahmed SYM, Cole A, Dawoud D, et al. Defining early health technology assessment: building consensus using Delphi technique. International Journal of Technology Assessment in Health Care. 2025;41(1):e34. 10.1017/S0266462325100123

14. Jorm A. Using the Delphi Method to Establish Expert Consensus: A Practical Guide: Springer Nature; 2025. 10.1007/978-981-96-8357-4

15. Okoli C, Pawlowski SD. The Delphi method as a research tool: an example, design considerations and applications. Information & Management. 2004;42(1):15–29. 10.1016/j.im.2003.11.002

16. Mchunu, G. (2012). Proposed guidelines for a workplace health promotion policy and implementation framework. Occupational Health Southern Africa, 18(2), 5–12. https://hdl.handle.net/10520/EJC119736

17. Hennink MM, Kaiser BN, Marconi VC. Code Saturation Versus Meaning Saturation:How Many Interviews Are Enough? Qualitative Health Research. 2017;27(4):591–608. 10.1177/1049732316665344

18. Ministry of Education, Ministry of Health. Kenya School Health Policy. 2018. https://www.ncikenya.or.ke/documents/KENYA%20SCHOOL%20HEALTH%20POLICY%20BOOK%20_20_1 1_2018.pdf

19. Ministry of Education, Ministry of Health, Ministry of Water Sanitation and Irrigation. Kenya Water Sanitation and Hygiene (WASH) in schools Strategy. Ministry of Health 2024. http://guidelines.health.go.ke/#/category/16/536/meta

20. United Nations Children’s Fund, World Health Organization. Progress on drinking water, sanitation and hygiene in schools: 2000-2021 Data update 2022 [Available from: https://data.unicef.org/resources/jmp-wash-in-schools-2022/.

21. United Nations Children’s Fund, World Health Organization. Progress on household drinking water, sanitation and hygiene 2000–2022: Special focus on gender 2023 [Available from: https://washdata.org/reports/jmp-2023-wash-households

22. Bohnert K, Chard AN, Mwaki A, Kirby AE, Muga R, Nagel CL, et al. Comparing Sanitation Delivery Modalities in Urban Informal Settlement Schools: A Randomized Trial in Nairobi, Kenya. International Journal of Environmental Research and Public Health. 2016;13(12):1189. https://www.mdpi.com/1660-4601/13/12/1189

23. Bah A, Diallo A, Bah A, Li F. Water, sanitation, and hygiene (WASH) coverage and practices of children from five public primary schools in Guinea. Journal of Water, Sanitation and Hygiene for Development. 2022;12(7):555–62. 10.2166/washdev.2022.078

24. Abdoulaye AA, Kadjangaba E, Léontine T, Bongo D, Jean-Claude DM. Evaluation of Quality Borehole Water Consumed in Public Schools in N’Djamena City (Chad). Journal of environmental science and engineering A II (2022). 2022:149–61.

25. Odeku OK. Critical Analysis of School Pit Toilet System as an Impediment to the Right to Access Quality Education in South Africa. African Journal of Public Affairs. 2022;13(1):97–109. doi:10.10520/ejc-ajpa_v13_n1_a6

26. Byansi JZ, Semiyaga S, Kansiime F, Kulabako RN. Factors enhancing operation and maintenance of sanitation facilities for improved service levels in Kampala Schools. BMC Public Health. 2025;25(1):1–16. 10.1186/s12889-025-21789-2

27. Azupogo UW, Dassah E, Bisung E. Promoting safe and inclusive water and sanitation services for students with physical disabilities in primary schools: a concept mapping study in Ghana. Journal of Water, Sanitation and Hygiene for Development. 2023;13(6):453–63. 10.2166/washdev.2023.029

28. Dassah E, Bisung E. Access to water, sanitation and hygiene (WASH) for persons with disabilities in school settings: A call for research. Disability & Society. 2023;38(5):893–8. 10.1080/09687599.2023.2181770

29. Wilbur J, Dreibelbis R, Mactaggart I. Addressing water, sanitation and hygiene inequalities: A review of evidence, gaps, and recommendations for disability-inclusive WASH by 2030. PLOS Water. 2024;3(6):e0000257. 10.1371/journal.pwat.0000257

30. Mukamana O, Johri M. What is known about school-based interventions for health promotion and their impact in developing countries? A scoping review of the literature. Health Education Research. 2016;31(5):587–602. https://search.ebscohost.com/login.aspx?direct=true&AuthType=sso&db=ccm&AN=118323066&site=e host-live&scope=site&custid=s5210036

31. McMichael C. Water, Sanitation and Hygiene (WASH) in Schools in Low-Income Countries: A Review of Evidence of Impact. Int J Environ Res Public Health. 2019;16(3). 10.3390/ijerph16030359

32. International Rescue Committe. WASH schools: Learning from the best 2020 [Available from: https://www.ircwash.org/news/wash-schools-learning-winners.

33. Riang’a RM, Nyanja N, Lusambili A, Mwangi EM, Ehrlich JR, Clyde P, et al. Implementation framework for income generating activities identified by community health volunteers (CHVs): a strategy to reduce attrition rate in Kilifi County, Kenya. BMC Health Services Research. 2024;24(1):132. https://link.springer.com/article/10.1186/s12913-023-10514-7

